# Lung Cancer Incidence in Counties at Low and High Risk of Radon Exposure: A Population-Based SEER Analysis (1975-2022)

**DOI:** 10.1101/2025.08.07.25333180

**Authors:** Nicholas Maxfield, Jason Semprini

**Author notes:** **Corresponding Author:** Jason Semprini 8026 Grand Ave West Des Moines, IA 50266.

## Abstract

**Background:** Lung cancer is the leading cause of cancer-related mortality in the United States. While smoking cigarettes remains the dominant risk factor, declining smoking rates have drawn attention to Radon. Despite the known risks from individual exposure to radon, less is known about long-term trends in Radon-related lung cancer at the population-level.

**Methods:** Using SEER-8 cancer registry data from four states, we compared lung cancer incidence between counties classified as low-risk and high-risk (>4.0 pCi/L) for radon exposure. Constructing a random effects Generalized Least Squares regression, which adjusted for county-level smoking prevalence, we quantified the excess lung cancer rate attributable to radon risk across decade, sex, and histologic subtype.

**Results:** Compared to low-risk counties, counties at high risk of radon exposure had a significantly higher overall lung cancer incidence: 13.5+ cases per 100,000 person-years (95% CI: 10.0, 17.1). This gap between low and high-risk counties was largest for Adenocarcinoma and small cell carcinoma, and larger in males than females. Only in females did we observe the gap in lung cancer incidence between low- and high-risk counties grow overtime.

**Conclusions:** This study illustrates how county-level radon data can be leveraged to enhance cancer surveillance and guide prevention and control strategies.

**Key Messages:**

- Radon is the second leading cause of lung cancer, and the primary cause among non-smokers.
- High-risk Radon counties had higher incidence of lung cancer over a nearly 50-year period.
- In females only, the gap between low and high-risk Radon counties grew decade after decade.

## Introduction

Lung cancer kills nearly 125,000 Americans annually[1]. Cigarette smoking contributes the most to lung cancer incidence[2]. However, smoking rates reached historic lows and continue to decline[3,4]. As a result, lung cancer incidence and mortality have declined dramatically[5]. Predictably, given that males smoke more frequently and at higher intensity than females, the observed 30-year decline in lung cancer for males (-38%) was remarkably greater than the stagnate trends in females (-1%)[5–8]. As smoking rates continue declining through the next decade, public health systems have the opportunity to prioritize secondary risk factors in the fight against lung cancer, which, despite 30-year declines, remains the leading cause of cancer mortality in the United States[1,9].

Second only to smoking, and first among non-smokers, the leading risk factor for lung cancer is Radon[10]. Radon has historically accounted for 10-15% of all lung cancers, but unlike the well known risk from smoking, the general public as well as clinicians appear unaware of the risks from Radon[10–12]. A naturally occurring radioactive gas formed by decaying uranium and radium, Radon seeps into buildings from the environment (i.e., soil, rocks, ground)[10]. Radon itself is harmless to humans, but its decay emits radioactive particles capable of attaching to dust, entering our lungs, and damaging our DNA[10,13–15].

In 1988, the International Agency for Research on Cancer classified Radon as carcinogenic to humans[16]. Around this same time in the United States, the Environmental Protection Agency (EPA) ramped up efforts to measure Radon across the country with the goal of understanding Radon’s impact on health[17]. In 1992, an EPA report identified a dose-response relationship between exposure to radon and lung cancer risk, as well as a synergistic relationship in people with a history of smoking[18]. Radon’s negative impact on health, apparently specific only to lung cancer, has since been observed consistently in various settings and communities[19–24].

## Objective

For nearly forty years, the scientific community has known that exposure to higher levels of Radon poses a risk to individuals. Yet, in our fight against lung cancer, two critical evidence gaps remain. First, most of the research since the 1988 classification has focused on individual-level risk of developing lung cancer after exposure to Radon as opposed to population-level surveillance. While individual-level research helps us understand and quantify the risk, population-level surveillance can directly inform public health system strategies. The one population-based study which analyzed Radon and lung cancer, included but a single state and just twenty years of data[23]. Second, while we know lung cancer incidence is declining as smoking rates decline, we know very little about changing trends in lung cancer attributable to Radon. One study attempted to investigate this trend, but failed to account for smoking prevalence and relied on broad national Radon level average; both of which appear inappropriate for understanding Radon attributable lung cancer trends in the United States[25].

Our study aimed to address these evidence gaps and examine the relationship between residential radon exposure and lung cancer incidence by sex, histologic subtype, and decade over a nearly 50-year time period. We hoped to inform public health strategies balancing smoking cessation and radon mitigation efforts in high-risk Radon areas.

## Methods

### Data and Measures

We analyzed data from the Surveillance, Epidemiology and End-Results (SEER-8) program (1975-2022)[26]. SEER-8 is a population-based cancer dataset of the original eight SEER cancer registries. For this study, we restricted our analysis to full state registries in the contiguous United States (Connecticut, Iowa, New Mexico, Utah). All cancer incidence rates were adjusted for age and analyzed at the county level. We further restricted our analysis to lung cancers (Lung and Bronchus). All rates were reported as cases per 100,000 person-years. In addition to reporting overall rates, we stratified the rates by sex (male, female), histologic subtype (Adenocarcinomas ICD-O-3: 8140-8389; Squamous cell carcinomas ICD-O-3: 8050; Small cell carcinomas ICD-O-3: 8043/3, 8044/3, 8045/3; and other), and decade of diagnosis (1975-1984, 1985-1994, 1995-2004, 2005-2014, 2015-2022). All counties were then classified into two groups, based on the EPA’s Map of Radon Zones[27]. This map categorizes U.S. counties based on radon measurements, geology, radioactivity, soil, and housing data. The EPA has established Radon levels above 4 pCi/L as high risk for developing lung cancer[28]. Therefore, we classified counties as “high-risk” if they exceeded the EPA’s mitigation threshold of 4.0. All other counties below 4.0 pCi/L were classified as “low-risk”.

### Statistical Analysis

To quantify the difference in lung cancer incidence rates between low and high-risk counties, we constructed a series of Random Effects Generalized Least Squares regression models[29–31]. Each model included a year fixed-effect, which adjusts for unobserved temporal trends in lung cancer incidence consistent across all counties. Additionally, each model included a county-level Random Effect, which adjusts for time-invariant differences between counties. Each model also adjusted for time-invariant county-level smoking differences [32]. For unbiased estimation, we assumed that the Random Effects component was uncorrelated with the other county level regressors. For inference, we estimated standard errors robust to autocorrelation and heteroskedasticity[33,34].

Equation 1 specifies the model, where Y_it_ is the lung cancer incidence rate for county = i, in ten-year period = t. The county-specific Random Effect is u_i_ and the idiosyncratic error term is e^i^. B is the parameter of interest, estimating the independent difference in lung cancer incidence rate between low and high-risk counties.

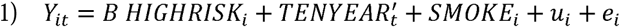

In addition to estimating overall differences between low and high-risk counties, we also tested if the difference changed over time. We modified Equation 1 by interacting the HIGHRISK variable with a set of decade (TENYEAR’) dummy variables.

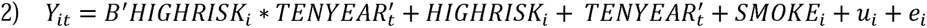

Now, Equation 2 estimated four B-parameters, each quantifying the decade-specific difference in lung cancer between low and high-risk counties relative to the difference in the reference decade (1975-1984). Finally, in addition to comparing confidence intervals of the estimates, we constructed a joint Wald test to test if the difference between low and high-risk counties changed over time. We used the Sidak correction method to adjust for multiple hypothesis tests (4 lung cancer subtypes x 4 decade tests x 3 groups), setting our alpha (a) for determining statistical significance at 1-(1-0.05)^(1/48) = 0.001[35].

## Results

### Summary Statistics

Table 1 shows the summary statistics. In the four states included in the analysis (1975-2022), 277,843 lung cancer cases were diagnosed. This corresponded to an ageladjusted incidence rate of 56.2 per 100,000 personlyears. 69% of the cases were diagnosed in counties with high-risk levels of Radon. The incidence rate in low-risk counties was 45.5 cases per 100,000 person-years and the incidence rate in high-risk counties was 60.0 cases per 100,000 person-years. Lung cancer incidence was highest in the states of Connecticut and Iowa, the decades of 1985-1994 and 1995-2004, Adenocarcinoma subtypes, and males.

**Table 1:** Summary Statistics. Reports the summary statistics of lung cancer incidence case data from SEER-8 (1975-2022). Counties were classified as high-risk of Radon from EPA classification (> 4pCi/L)

|  | **Count** | **(%)** | **Person-Years** | **Age-Adjusted Incidence Rate** |
| --- | --- | --- | --- | --- |
| **Overall** | 277,843 | (100) | 494,731,182 | 56.2 |
| **Low Risk Counties** | 86,090 | (31.0) | 208,761,159 | 45.4 |
| **High Risk Counties** | 191,753 | (69.0) | 285,970,023 | 60.0 |
| **Connecticut** | 115,584 | (41.6) | 162,472,669 | 63.9 |
| **Iowa** | 102,786 | (37.0) | 141,954,303 | 63.0 |
| **New Mexico** | 36,446 | (13.1) | 83,656,685 | 43.4 |
| **Utah** | 23,256 | (8.4) | 107,026,598 | 28.9 |
| **1975-1984** | 41,003 | (14.8) | 87,455,426 | 51.7 |
| **1985-1994** | 53,939 | (19.4) | 93,711,633 | 59.9 |
| **1995-2004** | 60,939 | (21.9) | 103,520,489 | 59.5 |
| **2005-2014** | 66,833 | (24.1) | 113,730,627 | 55.2 |
| **2015-2022** | 55,358 | (19.9) | 96,692,080 | 47.5 |
| **Adenocarcinoma** | 95,969 | (34.5) | 495,110,255 | 18.8 |
| **Squamous cell carcinoma** | 61,868 | (22.3) | 495,110,255 | 12.1 |
| **Small cell carcinoma** | 39,300 | (14.1) | 495,110,255 | 7.6 |
| **Other** | 80,935 | (29.1) | 495,110,255 | 16.0 |
| **Female** | 116,941 | (42.1) | 251,520,686 | 41.5 |
| **Male** | 161,131 | (58.0) | 243,589,569 | 71.8 |

### Overall Differences in Lung Cancer Incidence

Overall, after adjusting for smoking rate differences and county-level Random Effects, we estimated that high-risk counties were associated with an additional 13.5 lung cancer cases per 100,000 person-years (CI = 10.0, 17.1] (Table 2). Although each decade specific association was larger than the difference in 1975-1984, peaking in 2015-2022 (Est. = 13.1 cases per 100,000 person-years; CI = 9.0, 17.3), the associations did not vary significantly across decades (Wald Test Statistic p = 0.014). Figure 1 shows the decade-by-decade trends in lung cancer incidence. These overall differences were consistent across histological subtype (Table 2). However, the estimated association between high-risk counties and lung cancer incidence only varied across decades for Adenocarcinoma (Wald Test Statistic p <0.001), again peaking in 2015-2022 (Est. = 7.5 cases per 100,000 person-years, CI = 5.6, 9.5). Figure 2 shows the decade-by-decade trends in lung cancer incidence by histological subtype.

**Figure 1:**
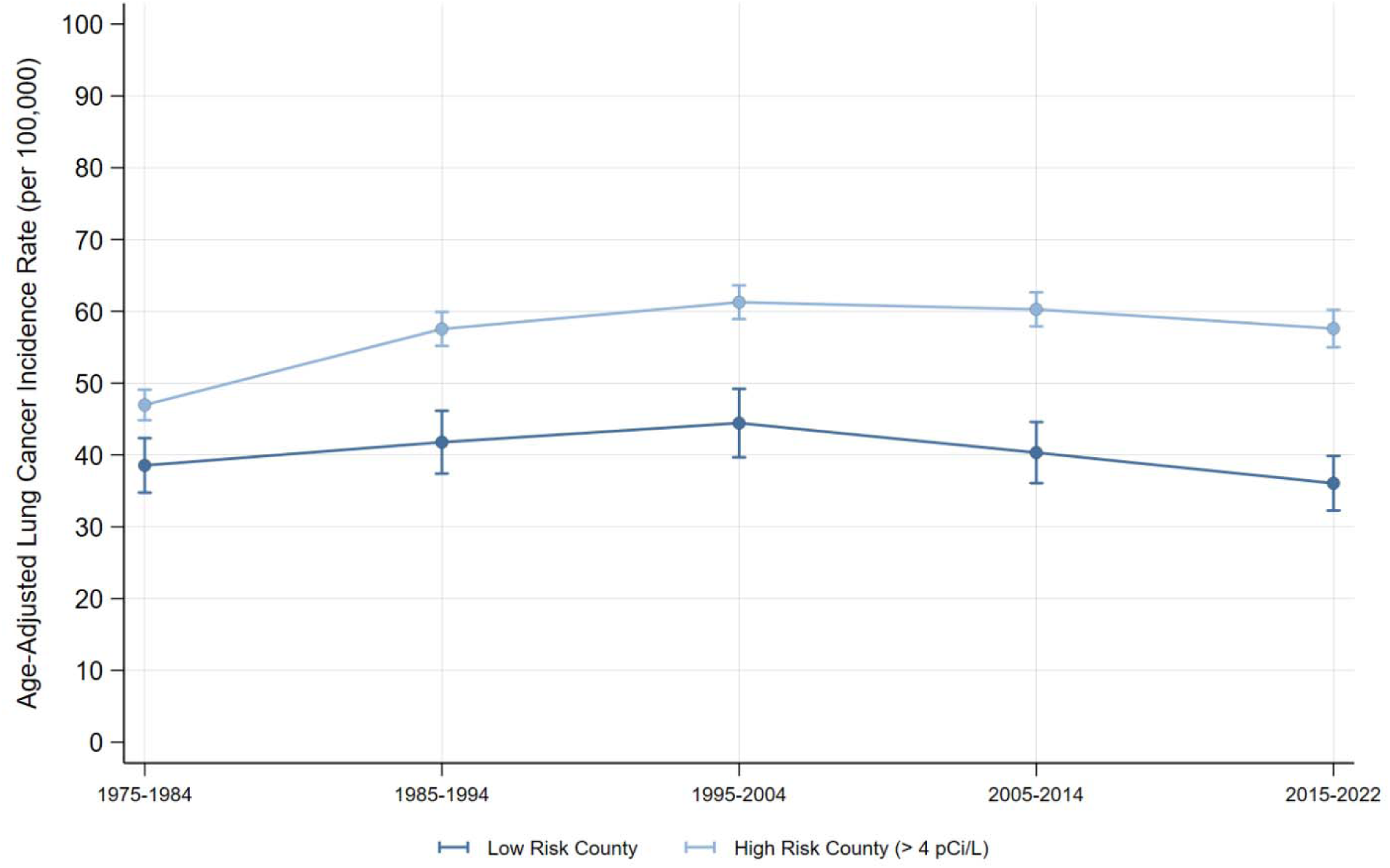
Lung Cancer Incidence (all subtypes) by Radon Exposure Risk. Visualizes the age-adjusted incidence of lung cancer in low and high-risk Radon counties. Source: SEER-8.

**Figure 2:**
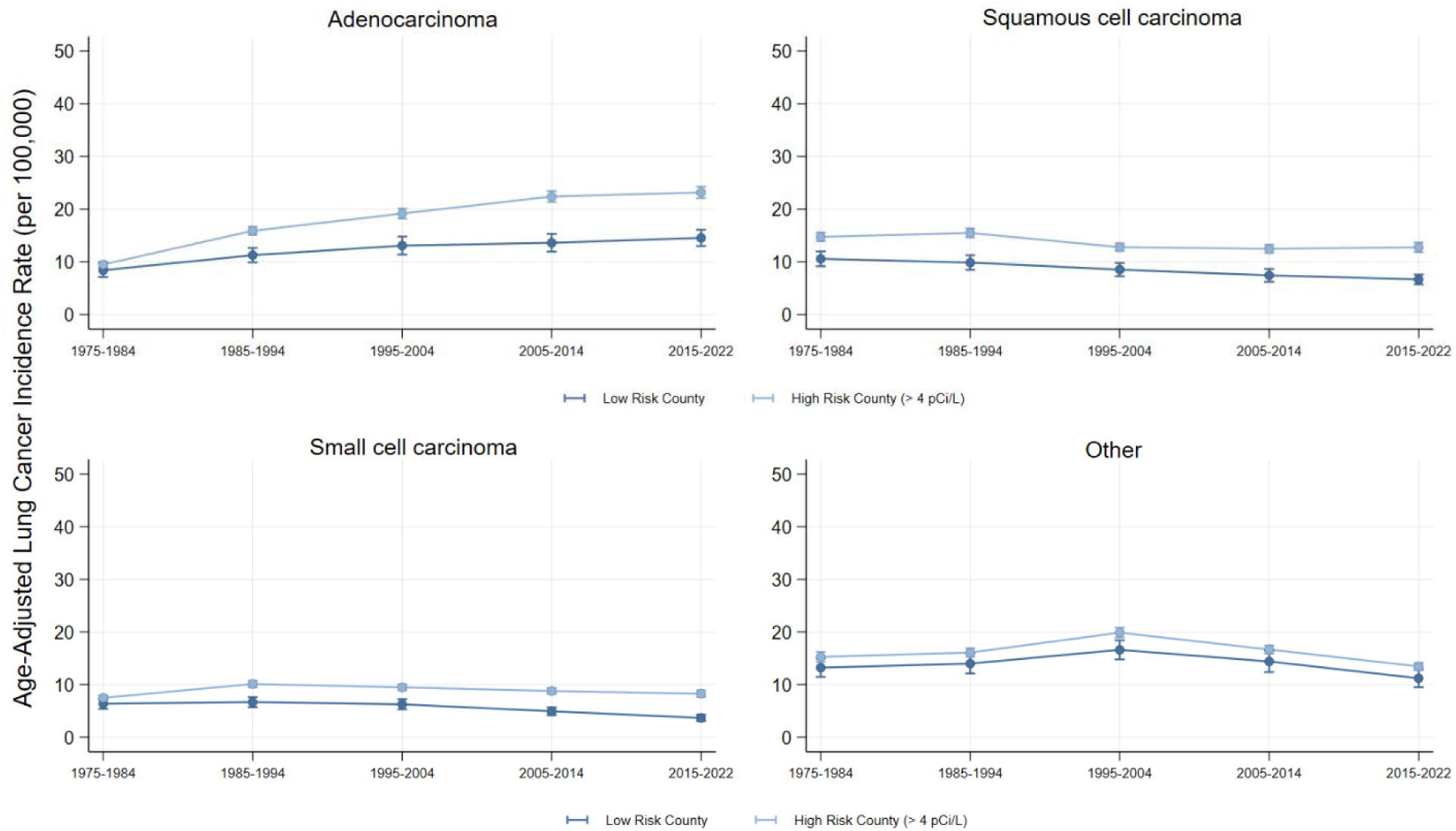
Lung Cancer Incidence by Radon Exposure Risk and Histologic Subtype. Visualizes the age-adjusted incidence of lung cancer in low and high-risk Radon counties, by histologic subtype. Source: SEER-8.

**Table 2:** Results from Random Effects GLS Regression Testing for Differences in Lung Cancer Incidence by County-Level Radon Risk (1975-2022) Reports the results of the Generalized Least Squares (GLS) Random Effects regression model equations. All sex groups and histologic subtypes were estimated separately. The first row of each sex-specific result estimated the association between High-Risk Radon County (> 4 pCi/L) and lung cancer incidence rate. 95% confidence intervals reported in brackets. The second set of estimates, estimated simultaneously, interact the High-Risk County parameter with a decade-specific parameter, estimating the relative difference between low and high-risk counties in that decade compared to the difference in 1975-1984. The Wald Test Statistic (p-value) tests whether the estimated associations varied across each decade. With the Sidak method, Wald Test Statistic alpha significance level = 0.001. All rates were adjusted for age and reported as cases per 100,000 person-years. * p <0.05, ** p <0.01, *** p <0.001.

|  | **Females and Males** |  |  |  |  |
| --- | --- | --- | --- | --- | --- |
| **Coefficient** | **All Subtypes** | **Adenocarcinoma** | **Squamous Cell** | **Small cell** | **Other** |
| High Risk County | 13.5***<br>[10.0, 17.1] | 5.2***<br>[4.0, 6.4] | 4.3***<br>[3.1, 5.4] | 2.7***<br>[-0.4, 1.7] | 1.4*<br>[0.0, 2.7] |
| High Risk X 1985-1994 | 7.3***<br>[3.7, 11.0] | 3.5***<br>[1.8, 5.3] | 1.5<br>[-0.2, 3.1] | 2.3***<br>[1.2, 3.4] | 0.1<br>[-1.7, 1.8] |
| High Risk X 1995-2004 | 8.4***<br>[4.7, 12.1] | 5.0***<br>[3.0, 7.0] | 0.1<br>[-1.5, 1.7] | 2.1***<br>[0.9, 3.3] | 1.2<br>[-0.6, 3.1] |
| High Risk X 2005-2014 | 11.5***<br>[7.8, 15.3] | 7.7***<br>[5.5, 10.0] | 0.9<br>[-0.5, 2.2] | 2.7***<br>[1.5, 3.9] | 0.2<br>[-1.6, 2.1] |
| High Risk X 2015-2022 | 13.1***<br>[9.0, 17.3] | 7.5***<br>[5.6, 9.5] | 1.9*<br>[0.4, 3.4] | 3.5***<br>[2.2, 4.7] | 0.2<br>[-2.0, 2.4] |
| Wald Test Statistic (p) | 0.014 | < 0.001 | 0.063 | 0.019 | 0.545 |
|  | **Females** |  |  |  |  |
| **Coefficient** | **All Subtypes** | **Adenocarcinoma** | **Squamous Cell** | **Small cell** | **Other** |
| High Risk County | 6.8***<br>[3.9, 9.6] | 3.6***<br>[2.4, 4.8] | 1.6***<br>[0.8, 2.3] | 1.7***<br>[1.1, 2.3] | -0.1<br>[-1.3, 0.4] |
| High Risk X 1985-1994 | 5.5**<br>[2.0, 8.9] | 2.9***<br>[1.3, 4.6] | 1.3<br>[-0.7, 3.2] | 1.5*<br>[0.3, 2.7] | -.2<br>[-1.8, 1.4] |
| High Risk X 1995-2004 | 9.3***<br>[5.4, 13.1] | 3.6**<br>[1.2, 6.0] | 1.4<br>[-0.2, 2.9] | 1.9*<br>[0.4, 3.5] | 2.4*<br>[0.4, 4.3] |
| High Risk X 2005-2014 | 15.0***<br>[10.9, 19.0] | 7.2***<br>[5.0, 9.5] | 3.3**<br>[2.3, 5.4] | 3.1***<br>[1.9, 4.3] | 1.3<br>[-0.8, 3.5] |
| High Risk X 2015-2022 | 18.5***<br>[14.5, 22.5] | 8.1***<br>[5.6, 10.5] | 3.9***<br>[1.8, 6.1] | 4.5***<br>[3.3, 5.6] | 2.1<br>[-0.8, 5.1] |
| Wald Test Statistic (p) | < 0.001 | < 0.001 | < 0.001 | < 0.001 | 0.025 |
|  | **Male** |  |  |  |  |
| **Coefficient** | **All Subtypes** | **Adenocarcinoma** | **Squamous Cell** | **Small cell** | **Other** |
| High Risk County | 23.5***<br>[18.5, 28.4] | 7.4***<br>[5.9, 8.9] | 8.1***<br>[6.3, 9.9] | 4.1***<br>[3.1, 5.1] | 3.9***<br>[2.0, 5.8] |
| High Risk X 1985-1994 | 10.1**<br>[3.9, 16.2] | 4.4*<br>[1.3, 7.5] | 1.9<br>[-0.7, 4.6] | 3.4**<br>[1.4, 5.5] | 0.3<br>[-2.9, 3.5] |
| High Risk X 1995-2004 | 6.4*<br>[0.3, 12.4] | 6.6***<br>[3.9, 9.3] | -2.0<br>[-4.9, 0.9] | 2.0*<br>[0.3, 3.8] | -.3<br>[-4.3, 1.6] |
| High Risk X 2005-2014 | 6.0<br>[-0.1, 12.2] | 8.0***<br>[4.8, 11.1] | -2.6<br>[-5.7, 0.5] | 2.1*<br>[0.2, 3.9] | -1.4<br>[-4.3, 1.6] |
| High Risk X 2015-2022 | 4.6<br>[-2.4, 11.5] | 6.7***<br>[3.7, 9.7] | -1.6<br>[-4.6, 1.5] | 2.3*<br>[0.2, 4.3] | -2.8<br>[-6.2, 0.6] |
| Wald Test Statistic (p) | 0.362 | 0.074 | 0.006 | 0.340 | 0.196 |

### Differences by Sex

#### Females

For females, we estimated that high-risk counties were associated with an additional 6.8 cases per 100,000 person-years (CI = 3.9, 9.6). Compared each of the three other histological subtypes, the association between high-risk counties and lung cancer incidence among females was larger in Adenocarcinomas (Est. = 3.6 cases per 100,000 person-years, CI = 2.4, 4.8). In fact, among females, there was no difference between low and high-risk counties for “other” lung cancer subtypes (Est. = -0.1 cases per 100,000 person-years, CI = -1.3, 0.4).

Overall and across the subtypes Adenocarcinoma, Squamous cell, and Small cell, the association between high-risk counties and lung cancer incidence among females grew significantly each decade (Wald Test Statistic p < 0.001). The largest association was for adenocarcinomas in 2015-2022 (Est. = 8.1 cases per 100,000 person-years, CI = 5.6, 10.5). See figures 3-4 for decade-by-decade trends of female lung cancer incidence overall and by subtype.

**Figure 3:**
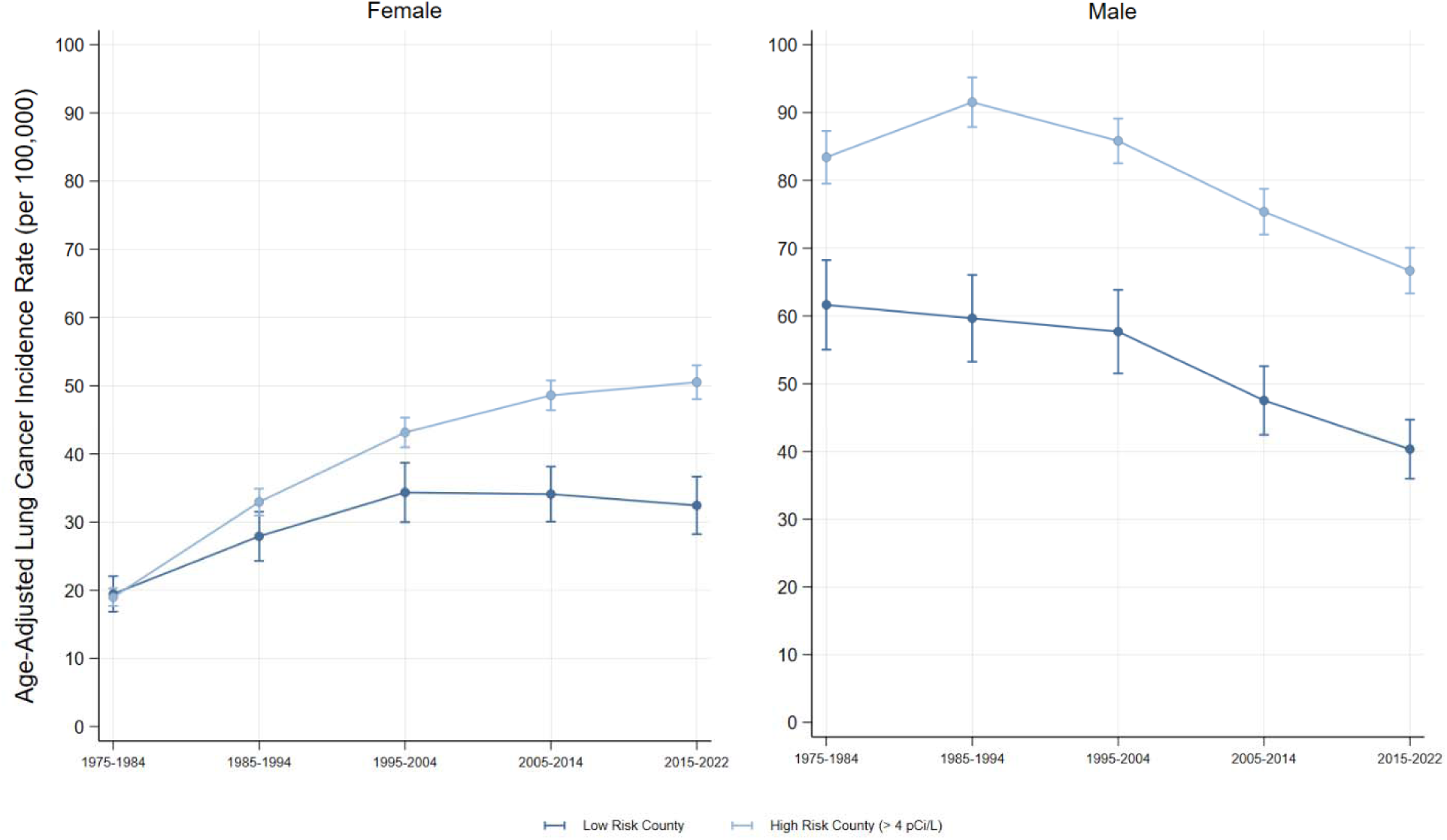
Lung Cancer Incidence (all subtypes) by Radon Exposure Risk and Sex. Visualizes the female and male age-adjusted incidence of lung cancer in low and high-risk Radon counties. Source: SEER-8.

**Figure 4:**
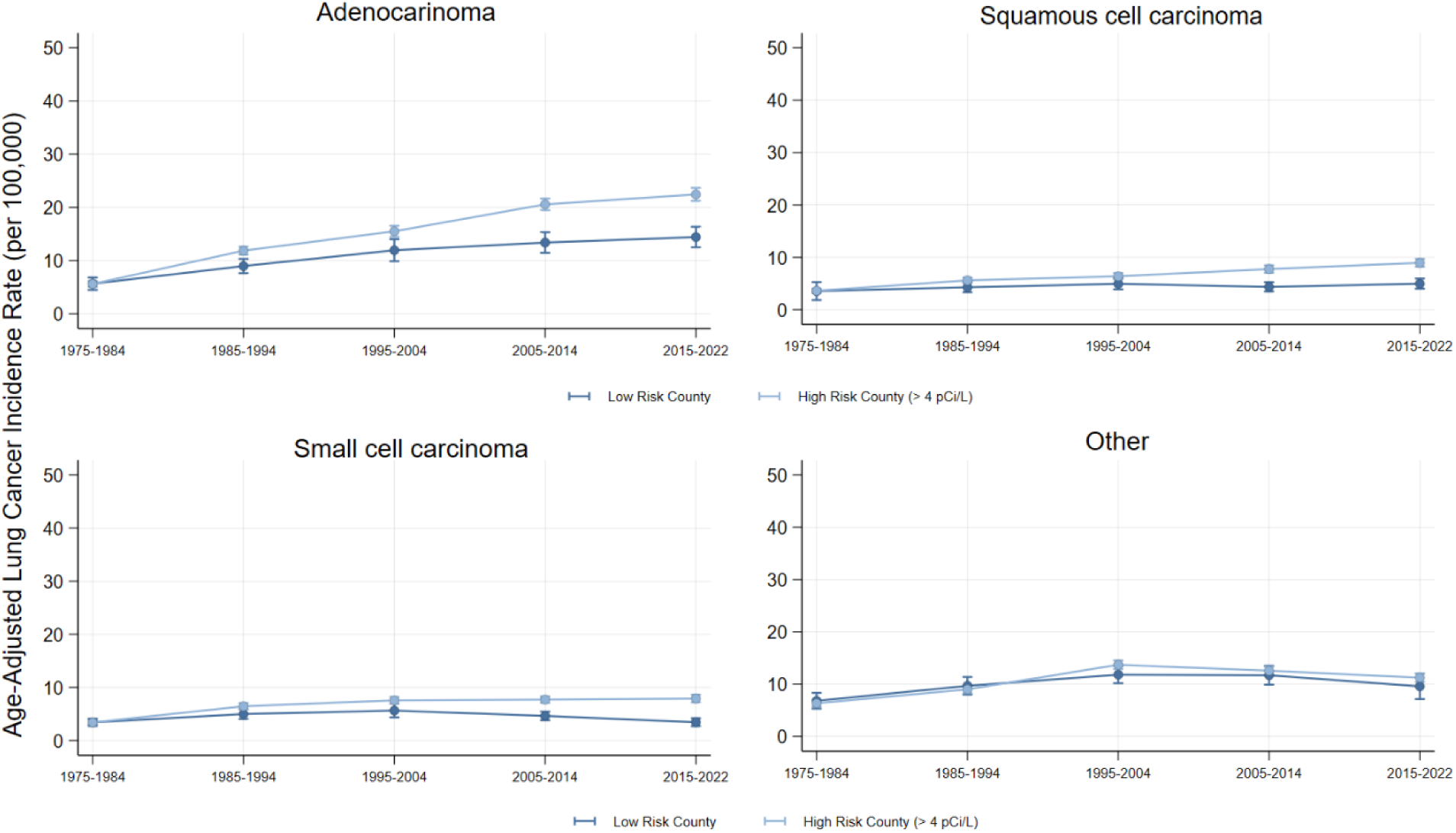
Female Lung Cancer Incidence by Radon Exposure Risk and Histologic Subtype. Visualizes the female age-adjusted incidence of lung cancer in low and high-risk Radon counties by histologic subtype. Source: SEER-8.

#### Males

In contrast, Males exhibited larger overall differences between low and high-risk counties, with an estimated association of 23.5 additional cases per 100,000 person-years (CI=18.5, 28.4). Compared to the “other” subtype, the associations were largest for adenocarcinoma and squamous cell carcinoma. Also contrary to the female estimates, the association between high-risk county and lung cancer incidence attenuated over time; where the 2005-2014 and 2015-2022 associations were no longer significantly different than the gap in 1975-1984. In fact, the only subtypes with statistically significant decade-specific coefficients were adenocarcinomas and small cell carcinomas; which again did not vary significantly across decades. See figures 3 and 5 for decade-by-decade trends of male lung cancer incidence overall and by subtype.

**Figure 5:**
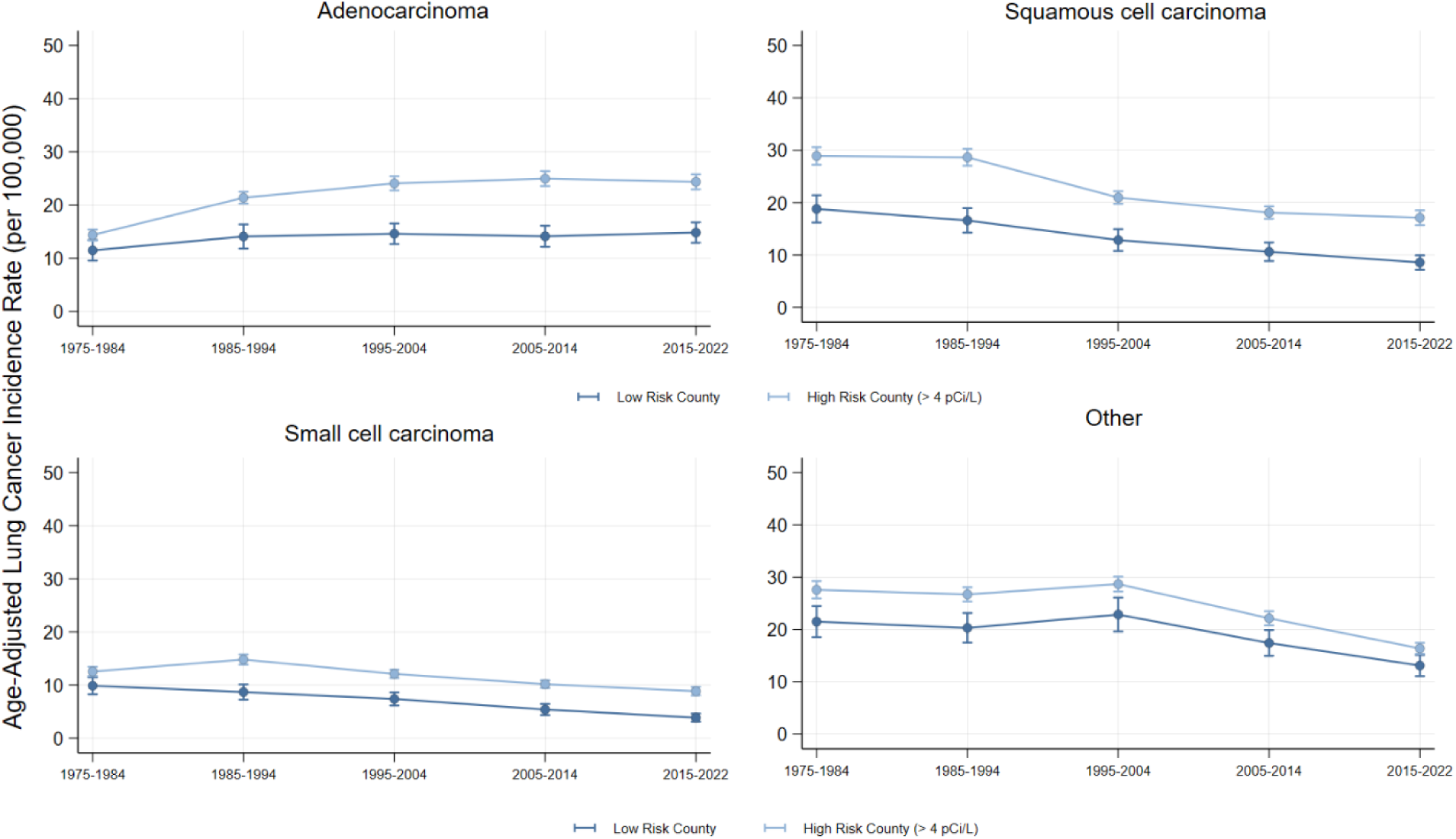
Male Lung Cancer Incidence by Radon Exposure Risk and Histologic Subtype. Visualizes the male age-adjusted incidence of lung cancer in low and high-risk Radon counties by histologic subtype. Source: SEER-8.

## Discussion

Our population-level analysis spanning nearly 50 years highlighted how lung cancer incidence trends varied by county-level of Radon risk. After adjusting for smoking, we observed large differences overall. This result validates the work of Ou, while also affirming that the elevated lung cancer rate in high Radon counties extends beyond the border of a single state[23]. The widening gap over time, however, was concentrated among Adenocarcinomas.

While this finding conflicts with previous research (which were notably not population-based, despite claiming to be so, and relied on point-in-time post-diagnosis Radon measures), our finding is supported by the data showing that 50-60% of the lung cancers diagnosed among never-smokers were adenocarcinomas[19,36,37].

Yet, the overall analyses masks sex-specific patterns in lung cancer incidence in high-risk Radon counties. Although the gap between low and high-risk counties was highest in males, that gap appeared to attenuate or stagnate over time. Whereas in females, for each of the three lung cancer subtypes, the gap between low and high-risk counties grew decade after decade. Further, despite only including four small states, the visual trends clearly mirror the overall trends in lung cancer incidence: dramatic declines in males, stagnate trends in females[5]. Since 1985-1994, we found each of the lung cancer types either declined or stagnated for both males and females, except female adenocarcinoma in high-risk Radon counties which has increased dramatically for half a century. Finally, by analyzing the most recent SEER data, we revealed that female lung cancer incidence in high-risk Radon counties now exceeds that of males in low-risk counties.

As smoking rates continue to decline, the proportion of lung cancers attributable to Radon will only grow. These high-risk Radon zones have been identified since 1993 and are unlikely to change, at least in terms of geological risk[27,28]. We cannot change what lies underneath the ground, we can only minimize how Radon emits into our homes, schools, and places of work. Notably, the EPA does not regulate Radon, but only provides education and guidance. States, rather, have been deemed responsible for protecting residents from the harm of Radon. Like most healthcare and public health policies, states have taken a wide range of approaches (testing, reporting, mitigation) in response to the threat of Radon[38,39].

Rather than conclude this study by examining the vast landscape of policy approaches taken, or not taken, we conclude this study by explicitly calling for novel areas of future research. We know Radon poses a significant health risk, potentially driving elevated rates of lung cancer in smokers and never smokers alike. We also know that policymakers have responded differently to this knowledge. What we do not yet know, unfortunately, is what works best at preventing Radon from emitting into homes, schools, and places of work. Neither do we know what works best at reducing downstream lung cancer risk among those with higher propensity of developing lung cancer in the future. To date, no research exists, to our knowledge, that has evaluated the causal impact of state or municipal policies on Radon emissions or lung cancer incidence. While simulation data exists in the U.S., real-world data may be more influential for policymakers[40,41]. This direction of scientific inquiry could involve retrospective analyses of policy changes over time or prospective evaluation (i.e., cluster randomized controlled trials). Such evidence would prove extremely valuable to state Medicaid programs, as well as federal and private health system payers, who pay the eventual cost of caring for patients diagnosed with lung cancer. This evidence would also provide a stronger tool for public health advocates, who currently rely solely on the need to reduce exposure to Radon, by providing concrete cost-effective solutions that advance population health and reduce healthcare expenditures.

Although Radon may be a growing driver of lung cancer, especially in never smokers, we would be remise to ignore the synergistic effect of Radon exposure among smokers[42]. The most recent data shows that 14% of American adults still smoke cigarettes, with higher rates among Black or Indigenous populations and people with underlying chronic conditions (i.e., disability, depression)[43]. While public health professionals should be celebrating the historic declines in cigarette smoking, we cannot afford to be complacent. Rather than solely balancing smoking cessation and Radon mitigation, public health systems should explore innovative approaches to combat both threats simultaneously. In fact, previous academic literature has shown the efficacy of leveraging perceived radon risk as a tool to inform the public about Radon and reduce smoking behavior[44–47]. Scaling these direct contact approaches with updated technology platforms across geographies and populations remains paramount[48–50]. States and municipalities could further extend synergistic harm reduction by building Radon mitigation into existing or new tobacco control initiatives (i.e., increasing cigarette taxes and spending the new revenue on Radon testing and mitigation support).

In 2013, Latz declared “Tobacco control policy is the most promising route to the public health goals of radon control policy”[51]. Since, the EPA, American Lung Association, and state of Kentucky have recently heeded the call[52–54]. As for the rest of us, the time to act is now. Because, over the next fifty years, we will be hard pressed to find any public health priority greater than eliminating the two main drivers of America’s deadliest cancer.

## Limitations

This study has several limitations. First, Radon exposure risk stratification may mask heterogeneity between counties within the high-risk group. We also, due to limited number of medium risk (2-4 pCi/L) counties, combined all counties not deemed high-risk. Second, Radon exposure was assessed at the county level, as was cancer incidence, which prohibited us from analyzing more granular geographic variation. Third, smoking data used as a control was limited to 2003-2011 and was not sex-stratified and did not account for intensity or duration. Fourth, the SEER-8 data includes four small states, which represent only 4% of the population. Application of our findings may only generalize to smaller and racially homogenous rural states. Finally, our analysis spanned a large time-period and did not attempt to adjust for dynamic policy landscapes related to smoking cessation or Radon mitigation.

Finally, like all observational research, the design was subject to internal validity threats given the potential for unobservable county-level factors may be correlated with both the county-level Random Effect and Radon risk group. For this reason, we avoided causal language and reported our results in terms of estimated associations.

## Conclusions

As smoking rates in the United States continue to decline, public health systems balance prioritizing smoking cessation efforts with preventing secondary causes of lung cancer such as Radon. This study aimed to evaluate the relationship between high-risk residential radon exposure and lung cancer incidence by sex, histologic subtype, and decade over a nearly 50-year time period. Overall, we found that counties with high Radon accounted for most of the lung cancer cases and had a significantly higher age adjusted incidence rate when compared to low risk Radon counties. Most concerning, we found that the gap in lung cancer incidence between low and high-risk Radon counties were rising for females, resulting in a reversal of rankings for females in high-risk counties compared to males in low-risk counties. The limited regulatory efforts by the EPA have left the authority over protecting the public from the harms of Radon has fallen to states and municipalities, ultimately resulting in a vast landscape of radon testing and mitigation policies in both low and high-risk regions. Regardless of policy approaches, public health systems have an opportunity to capitalize growing interest in smoking cessation and Radon mitigation by creating novel programs which support both priorities in pursuit of reducing lung cancer risk for all Americans.

## Data Availability

The SEER-8 datafile used in this analysis is included as a supplemental file.

## Notes

### Competing Interest Statement

The authors have declared no competing interest.

### Funding Statement

This study did not receive any funding.

### Summary of Updates

The following statement was removed from the first paragraph of the methods section, as it was redundant and incomplete: "County-level cigarette smoking rates are a critical factor for analyzing Radon and lung cancer. We accounted for".

